# Risk factors for experiencing Long-COVID symptoms: Insights from two nationally representative surveys

**DOI:** 10.1101/2024.01.12.24301170

**Authors:** Yixuan Wu, Mitsuaki Sawano, Yilun Wu, Rishi M. Shah, Pamela Bishop, Akiko Iwasaki, Harlan M. Krumholz

**Affiliations:** Department of Biostatistics, Yale School of Public Health, New Haven, Connecticut; Center for Outcomes Research and Evaluation, Yale New Haven Hospital, New Haven, Connecticut; Center for Infection and Immunity, Yale School of Medicine, New Haven, Connecticut; Department of Applied Mathematics, Yale College, New Haven, Connecticut; Department of Immunobiology, Yale School of Medicine, New Haven, Connecticut; Howard Hughes Medical Institute, Chevy Chase, Maryland; Section of Cardiovascular Medicine, Department of Internal Medicine, Yale School of Medicine, New Haven, Connecticut; Department of Health Policy and Management, Yale School of Public Health, New Haven, Connecticut

## Abstract

**Background:** Long COVID (LC) is a complex and multisystemic condition marked by a diverse range of symptoms, yet its associated risk factors remain poorly defined.

**Methods:** Leveraging data from the 2022 Behavioral Risk Factor Surveillance System (BRFSS) and National Health Interview Survey (NHIS), both representative of the United States population, this study aimed to identify demographic characteristics associated with LC. The sample was restricted to individuals aged 18 years and older who reported a positive COVID-19 test or doctor’s diagnosis. We performed a descriptive analysis comparing characteristics between participants with and without LC. Furthermore, we developed multivariate logistic regression models on demographic covariates that would have been valid at the time of the COVID-19 infection.

**Results:** Among the 124,313 individuals in BRFSS and 10,131 in the NHIS reporting either a positive test or doctor’s diagnosis for COVID-19 (Table), 26,783 (21.5%) in BRFSS and 1,797 (17.1%) in NHIS reported LC. In the multivariate logistic regression model, we found middle age, female gender, Hispanic ethnicity, lack of a college degree, and residence in non-metropolitan areas associated with higher risk of LC. Notably, the initial severity of acute COVID-19 was strongly associated with LC risk. In contrast, significantly lower ORs were reported for Non-Hispanic Asian and Black Americans compared to Non-Hispanic White.

**Conclusions:** In the United States, there is marked variation in the risk of LC by demographic factors and initial infection severity. Further research is needed to understand the underlying cause of these observations.

## Introduction

Long COVID (LC) is a multisystemic condition characterized by a diverse range of symptoms,^1^ and the risk factors associated with LC are not well defined. The most recent versions of the Behavioral Risk Factor Surveillance System (BRFSS) survey and National Health Interview Survey (NHIS) have sampling and survey designs suitable to determine the patient demographic characteristics associated with LC.^2^

## Methods

The study data are from the 2022 BRFSS and NHIS, which included individuals ≥18 years living in the United States. We restricted the sample to people who reported either a positive test or a doctor’s diagnosis of COVID-19. An affirmative or negative response to this question defined LC or non-LC, respectively: “Did you have any symptoms lasting 3 months or longer that you did not have before having coronavirus or COVID-19?”. The COVID-19 severity question is exclusive to the NHIS data, while other characteristics included in the study are consistent. We used chi-squared tests to compare characteristics between participants with and without LC. We developed multivariate logistic regression models on demographic covariates that would have been valid at the time of the COVID-19 infection (Table). We reported odds ratios (OR) to identify risk factors associated with LC. All analyses were performed using R, version 4.1.2 (R Foundation for Statistical Computing).

## Results

Among the 124,313 individuals in BRFSS and 10,131 in the NHIS reporting either a positive test or doctor’s diagnosis for COVID-19 (Table), 26,783 (21.5%) in BRFSS and 1,797 (17.1%) in NHIS reported LC. The sociodemographic characteristics of those reporting LC were largely consistent across both surveys (Table and Figure). Those with more severe acute COVID-19 presentations had the highest OR. In addition, older adults, females, people who identified as Hispanic, those without a college degree, and those living in non-metropolitan areas had higher risks of being affected by LC. Of note, the oldest adults (aged ≥ 65 years) had risks that were only slightly higher than those of the youngest adults (aged 18-24 years). Non-Hispanic Black and Asian people had lower risks than White people, with Asian people having the lowest risk.

**Table.**
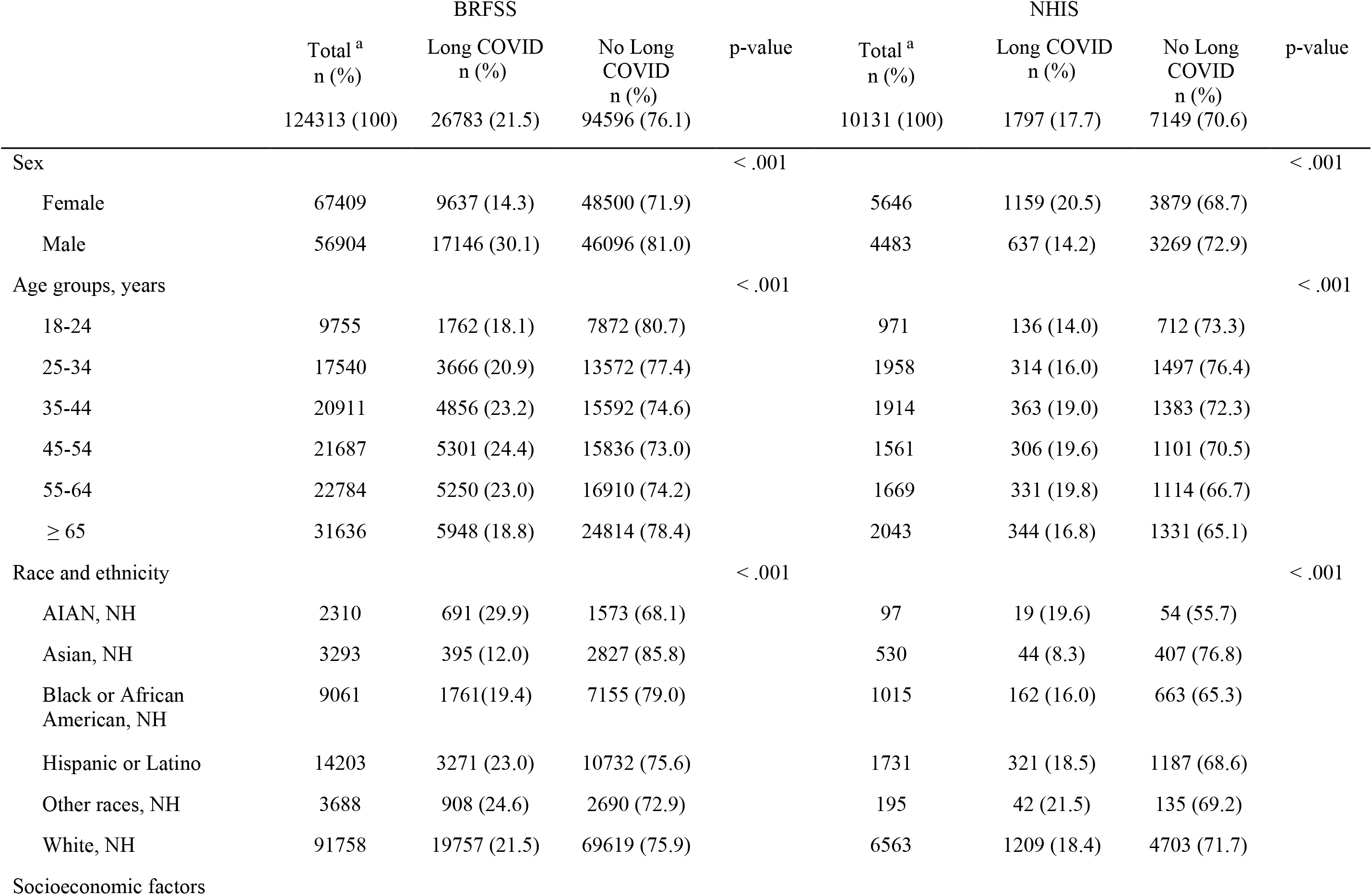
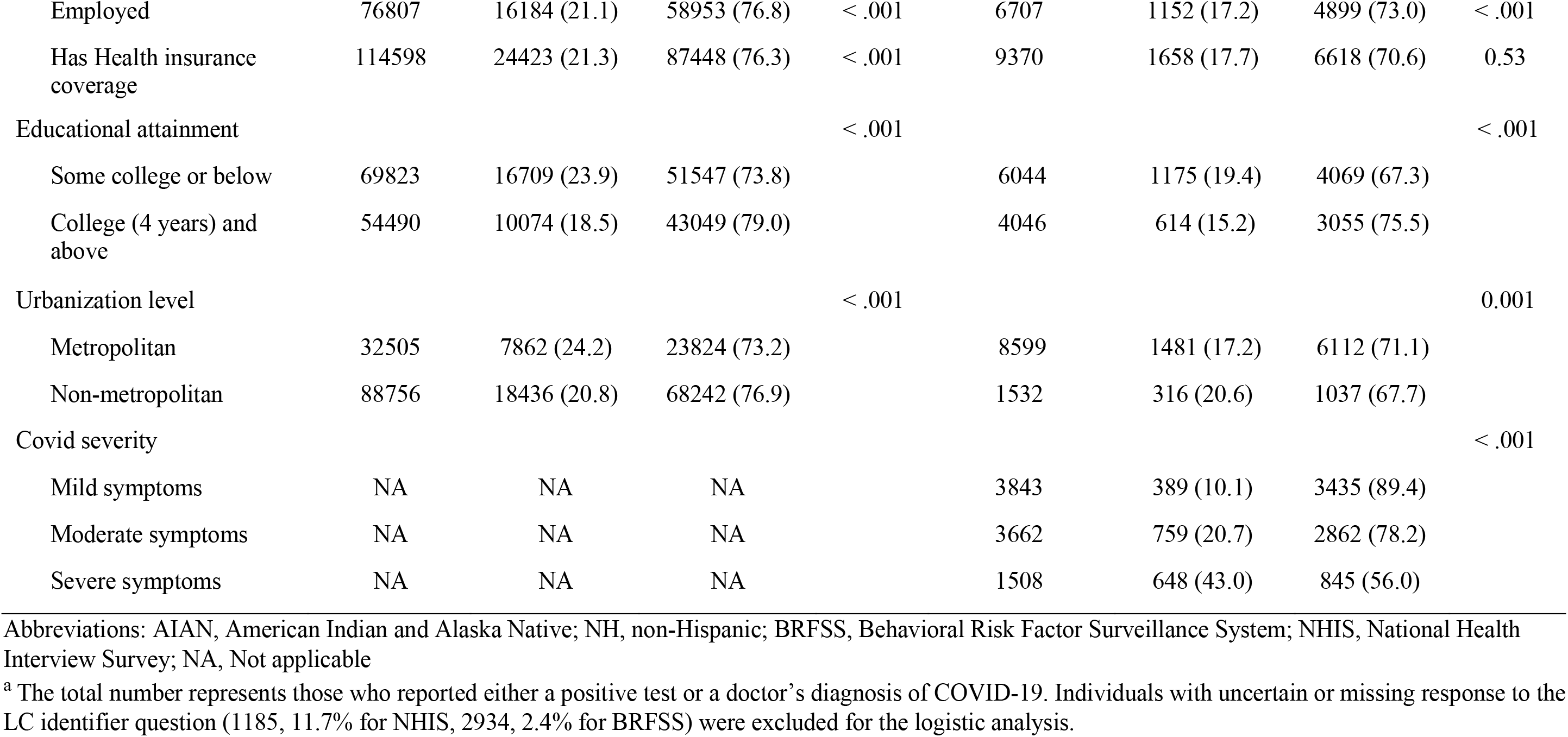
Sociodemographic characteristics of patients with and without Long Covid symptoms in the BRFSS and NHIS.

|  | BRFSS |  |  | p-value | NHIS |  |  | p-value |
| --- | --- | --- | --- | --- | --- | --- | --- | --- |
|  | Total <sup>a</sup><br>n (%) | Long COVID<br>n (%) | No Long<br>COVID<br>n (%) |  | Total <sup>a</sup><br>n (%) | Long COVID<br>n (%) | No Long<br>COVID<br>n (%) |  |
|  | 124313 (100) | 26783 (21.5) | 94596 (76.1) |  | 10131 (100) | 1797 (17.7) | 7149 (70.6) |  |
| Sex |  |  |  | < .001 |  |  |  | < .001 |
| Female | 67409 | 9637 (14.3) | 48500 (71.9) |  | 5646 | 1159 (20.5) | 3879 (68.7) |  |
| Male | 56904 | 17146 (30.1) | 46096 (81.0) |  | 4483 | 637 (14.2) | 3269 (72.9) |  |
| Age groups, years |  |  |  | < .001 |  |  |  | < .001 |
| 18-24 | 9755 | 1762 (18.1) | 7872 (80.7) |  | 971 | 136 (14.0) | 712 (73.3) |  |
| 25-34 | 17540 | 3666 (20.9) | 13572 (77.4) |  | 1958 | 314 (16.0) | 1497 (76.4) |  |
| 35-44 | 20911 | 4856 (23.2) | 15592 (74.6) |  | 1914 | 363 (19.0) | 1383 (72.3) |  |
| 45-54 | 21687 | 5301 (24.4) | 15836 (73.0) |  | 1561 | 306 (19.6) | 1101 (70.5) |  |
| 55-64 | 22784 | 5250 (23.0) | 16910 (74.2) |  | 1669 | 331 (19.8) | 1114 (66.7) |  |
| ≥ 65 | 31636 | 5948 (18.8) | 24814 (78.4) |  | 2043 | 344 (16.8) | 1331 (65.1) |  |
| Race and ethnicity |  |  |  | < .001 |  |  |  | < .001 |
| AIAN, NH | 2310 | 691 (29.9) | 1573 (68.1) |  | 97 | 19 (19.6) | 54 (55.7) |  |
| Asian, NH | 3293 | 395 (12.0) | 2827 (85.8) |  | 530 | 44 (8.3) | 407 (76.8) |  |
| Black or African<br>American, NH | 9061 | 1761(19.4) | 7155 (79.0) |  | 1015 | 162 (16.0) | 663 (65.3) |  |
| Hispanic or Latino | 14203 | 3271 (23.0) | 10732 (75.6) |  | 1731 | 321 (18.5) | 1187 (68.6) |  |
| Other races, NH | 3688 | 908 (24.6) | 2690 (72.9) |  | 195 | 42 (21.5) | 135 (69.2) |  |
| White, NH | 91758 | 19757 (21.5) | 69619 (75.9) |  | 6563 | 1209 (18.4) | 4703 (71.7) |  |
| Socioeconomic factors |  |  |  |  |  |  |  |  |
| Employed | 76807 | 16184 (21.1) | 58953 (76.8) | < .001 | 6707 | 1152 (17.2) | 4899 (73.0) | < .001 |
| Has Health insurance coverage | 114598 | 24423 (21.3) | 87448 (76.3) | < .001 | 9370 | 1658 (17.7) | 6618 (70.6) | 0.53 |
| Educational attainment |  |  |  | < .001 |  |  |  | < .001 |
| Some college or below | 69823 | 16709 (23.9) | 51547 (73.8) |  | 6044 | 1175 (19.4) | 4069 (67.3) |  |
| College (4 years) and above | 54490 | 10074 (18.5) | 43049 (79.0) |  | 4046 | 614 (15.2) | 3055 (75.5) |  |
| Urbanization level |  |  |  | < .001 |  |  |  | 0.001 |
| Metropolitan | 32505 | 7862 (24.2) | 23824 (73.2) |  | 8599 | 1481 (17.2) | 6112 (71.1) |  |
| Non-metropolitan | 88756 | 18436 (20.8) | 68242 (76.9) |  | 1532 | 316 (20.6) | 1037 (67.7) |  |
| Covid severity |  |  |  |  |  |  |  | < .001 |
| Mild symptoms | NA | NA | NA |  | 3843 | 389 (10.1) | 3435 (89.4) |  |
| Moderate symptoms | NA | NA | NA |  | 3662 | 759 (20.7) | 2862 (78.2) |  |
| Severe symptoms | NA | NA | NA |  | 1508 | 648 (43.0) | 845 (56.0) |  |

Abbreviations: AIAN, American Indian and Alaska Native; NH, non-Hispanic; BRFSS, Behavioral Risk Factor Surveillance System; NHIS, National Health Interview Survey; NA, Not applicable <sup>a</sup> The total number represents those who reported either a positive test or a doctor's diagnosis of COVID-19. Individuals with uncertain or missing response to the LC identifier question (1185, 11.7% for NHIS, 2934, 2.4% for BRFSS) were excluded for the logistic analysis.

**Figure.**
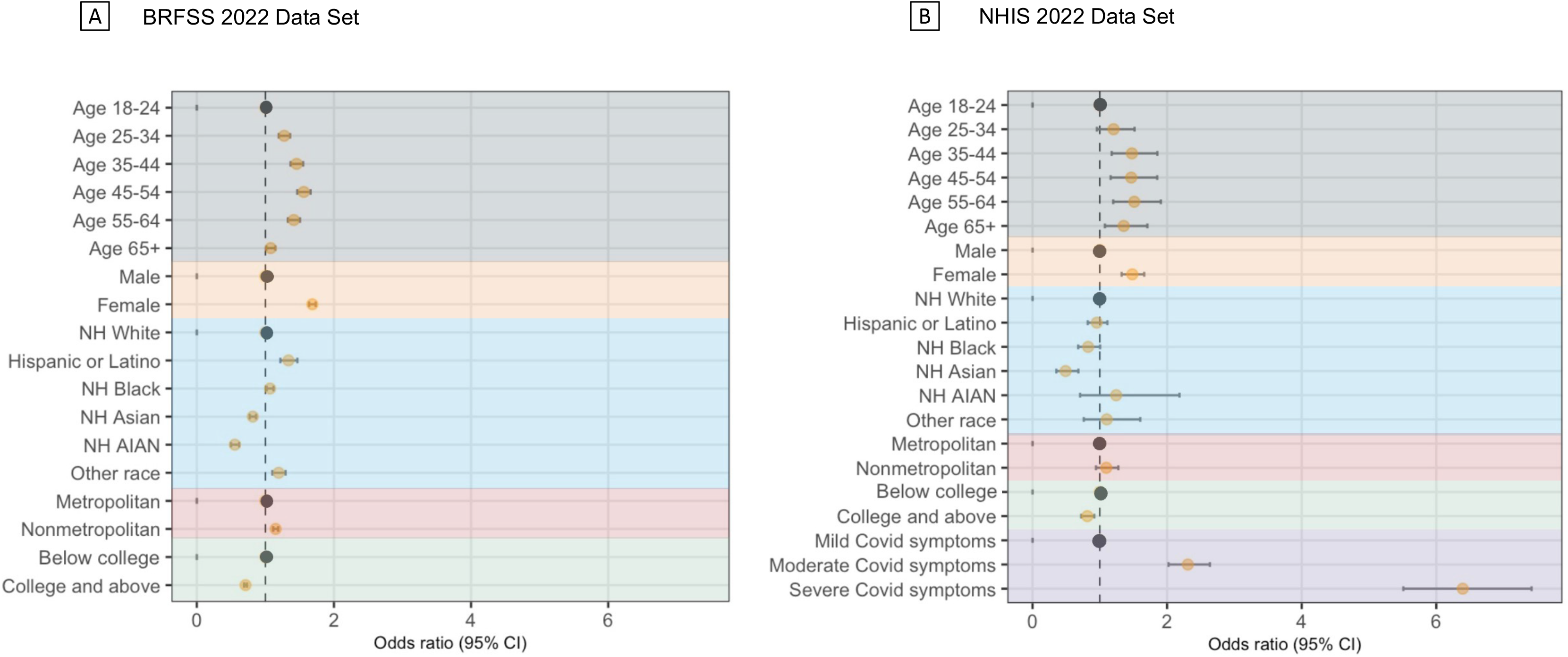
Risk factors and associations with Long-COVID symptoms in patients. Risk factors described by ORs (95% CI) of multivariate logistic regression for the NHIS data set (A) and BRFSS data set (B). Black dots represent the reference category for each variable. Individuals who had more severe covid symptoms had a significantly higher risk of experiencing LC symptoms. Abbreviations: AIAN, American Indian and Alaska Native; NH, non-Hispanic; BRFSS, Behavioral Risk Factor Surveillance System; NHIS, National Health Interview Survey

## Discussion

This study identified demographic characteristics associated with LC. We identified the initial severity of the infection as strongly associated with LC, which may point to prevention and early intervention with therapeutics as an important approach. The reason for the variation in the risks associated with demographic factors, such as sex, race and ethnicity, education, and geography, is unclear and could be related to exposures, treatments, and reporting. There is a need for further studies to illuminate the underlying mechanisms of these differences. Although this study is based on self-reported data, this is currently the only way to diagnose LC, and we restricted the sample to those who reported either a positive test or doctor’s diagnosis for COVID-19. A strength of this study is the consistency in these national surveys. Other studies have reported female sex, older age, and previous hospitalization or ICU admission in association with higher risk for LC.^3-5^ However, previous studies did not have the advantage of national representation from a single country that experienced a substantial pandemic toll. Limitations of our study include the lack of medical record information, though that information would be incomplete for many people. In conclusion, in the United States, there is marked variation in the risk of LC by demographic factors and initial infection severity. Further research is needed to understand the underlying cause of these observations.

## Data Availability

All data produced are available online at CDC official website.

## References

1. Davis HE, McCorkell L, Vogel JM, Topol EJ. Long COVID: major findings, mechanisms and recommendations [published correction appears in Nat Rev Microbiol. 2023 Jun;21(6):408]. Nat Rev Microbiol. 2023;21(3):133–146. doi:10.1038/s41579-022-00846-2

2. Adjaye-Gbewonyo D, Vahratian A, Perrine CG, Bertolli J. Long COVID in Adults: United States, 2022. NCHS Data Brief. Sep 2023;(480):1–8.

3. Tsampasian V, Elghazaly H, Chattopadhyay R, et al. Risk factors associated with post-COVID-19 condition: a systematic review and meta-analysis. JAMA Intern Med. 2023;183(6):566–580. doi:10.1001/jamainternmed.2023.0750

4. Bai F, Tomasoni D, Falcinella C, et al. Female gender is associated with long COVID syndrome: a prospective cohort study. Clin Microbiol Infect. 2022;28(4):611.e9-611.e16. doi:10.1016/j.cmi.2021.11.002

5. Sudre CH, Murray B, Varsavsky T, et al. Attributes and predictors of long COVID [published correction appears in Nat Med. 2021 Jun;27(6):1116]. Nat Med. 2021;27(4):626–631. doi:10.1038/s41591-021-01292-y

